# Rare case of Pediatric Male Secretory Breast Carcinoma: An Integrative study with Genomic and Transcriptomic analysis

**DOI:** 10.1101/2025.11.20.25337809

**Authors:** Sohini Guha, Somrita Das, Shashikumar T, Kiruthiga Raghunathan, Pragnya Coca, Shobha Badiger, Sujan K. Dhar, Manjula Das

## Abstract

**Background:** Male breast cancer constitutes <1% of all breast cancer cases and is exceedingly rare in children, with pediatric male secretory breast carcinoma (SBC) reported only as isolated case reports. SBC is typically triple-negative and indolent, driven by the characteristic ETV6-NTRK3 fusion. However, the molecular landscape of aggressive pediatric male SBC remains poorly understood.

**Objectives:** To characterise the genomic and transcriptomic features of a rare pediatric male case of SBC and identify clinically actionable molecular alterations using integrated next-generation sequencing.

**Methods:** Retrospective fresh frozen tumour material underwent RNA and DNA extraction followed by whole-exome sequencing (WES) and transcriptome profiling using Illumina HiSeq X (150 bp paired-end). RNA-seq data were analysed using fastp, STAR, STAR-Fusion, and DESeq2. Mutational annotation was performed with COSMIC. Expression profiles were compared with ERBB2-stratified male breast cancers from TCGA-BRCA. Immune checkpoint and fusion interactome analyses were conducted to identify activated pathways. No formal statistical testing was performed due to the single-patient nature of the study.

**Results:** WES identified fifteen somatic exonic mutations, including recurrent alterations in ACAD11, PTPRM, INSR, and OR6A2, implicating pathways involved in metabolism, cell adhesion, signalling, and inflammatory responses. Transcriptomic profiling revealed high expression of secretory lineage genes (SCGB2A2, SCGB1D2), stress-response mediators (FTH1, CLU, S100A9), and biosynthetic markers (ribosomal proteins, TMSB4X). Hormone receptors (ESR1, PGR, AR) were negligible, whereas ERBB2 exhibited appreciable expression, suggesting a HER2-enriched subtype atypical for SBC. Immune checkpoint analysis showed elevated HMGB1, LAG3, LGALS9, PD-L1, and CD86 expression. Fusion analysis identified the canonical ETV6-NTRK3 rearrangement with validated breakpoints and evidence of MAPK/ERK and PI3K/AKT pathway activation. HER2 overexpression in this post-treatment sample may be attributable to gemcitabine-induced NF-κB activation or initial false-negative IHC.

**Conclusion:** This integrative genomic-transcriptomic analysis uncovered multiple clinically actionable alterations in a rare pediatric male SBC, including HER2 expression and ETV6-NTRK3 fusion. These findings demonstrate the diagnostic and therapeutic value of comprehensive NGS in rare, aggressive paediatric tumours and highlight its potential to guide precision oncology. Early incorporation of transcriptomic profiling into diagnostic pathways may substantially improve treatment opportunities, particularly in resource-limited settings.

## Introduction

Male breast cancer accounts for < 1% of all breast cancer cases and 0.5% of all malignancies in men. [1,2] This malignancy can originate from the epithelium of the ducts or lobules of the breast. Male breast cancer risk factors include family history, radiation exposure, estrogen therapy, hyperestrogenic states (cirrhosis, Klinefelter’s), and testicular disorders such as undescended testes, orchitis, and infertility [3]. Analysis of a large cohort (n = 2,696,734) indicated the age at diagnosis to be ranging from 47 to 77 years, with mean age at 60 years [4,5]. Primary breast cancer in the pediatric population is exceedingly rare, representing <0.1% of all breast cancers and <1% of all pediatric malignancies [6,7]. While the vast majority of cases occur in females, male pediatric breast cancer is found in the literature primarily as isolated case reports. In a literature review of 32 cases (ages 4–17 years), 62.5% occurred in females and 37.5% in males [6]. Overall, pediatric male breast cancer remains an exceptional finding compared with the female counterpart.

Though the disease is rarer in the pediatric population, it appears more aggressive than in adults [8]. Secretory breast cancer (SBC) is a rare kind of breast cancer, accounting for 0.15% of all infiltrating breast cancers, with a male-to-female ratio of 1:31 according to National Cancer Database analysis [9]. According to the World Health Organisation (WHO), breast tumours are classified among exceptionally rare tumour types [10]. Secretory breast carcinoma (SBC) represents the most common breast cancer in children, yet it remains rare overall, comprising less than 1% of all childhood malignancies [11–14].

SBC is even rarer in males, with 15 reported cases until 2004 [15,16]. Previous studies indicate that SBC is negative for estrogen receptor (ER), progesterone receptor (PR), and human epidermal growth factor receptor 2 (HER2), but positive for basal-cell markers; therefore, it has been classified as a specific subtype of triple-negative breast cancer (TNBC). It is estimated that 10% of men with breast cancer have a genetic predisposition such as BRCA1, BRCA2, PTEN, P53, CHEK21, androgen receptor (AR) gene mutations or CYP17 polymorphism [17]. Moreover, it has been reported that there is an increased risk of cancer in 5-10% of men who carry mutations in the tumour suppressor gene BRCA2 [18,19].

In 2002, SBC was first reported to harbour the following recurrent balanced chromosomal translocation: t(12;15) (p13; q25). This translocation leads to the formation of the ETV6-NTRK3 fusion [20]. Pediatric SBC has been reported to have distinctive molecular features. In a previous study, a 6-year-old boy had a triple-negative tumour with basal and luminal markers, harbouring the classic t(12;15) fusion plus a 3q28 duplication (also present in his father and grandfather with prior breast cancer, but without BRCA mutations) [21]. Another case in a 5-year-old boy revealed an ETV6-NTRK3 translocation and PDGFRB c.2632A>G mutation, but no BRCA1/2, TP53, RAD51C, or RAD51D alterations [22]. Overall, such tumours show a basal-like phenotype (CK5/6 or EGFR positive), diffuse MUC4, frequent SOX10 positivity, and are triple-negative or weakly ER-positive [23].

Unlike basal/triple-negative breast cancers of no special type, secretory carcinomas exhibited an exceptionally low mutational burden, lacking pathogenic alterations in common cancer-related genes such as BRCA1, BRCA2, TP53, RAD51C, and RAD51D. [20] These findings highlight ETV6–NTRK3 as the principal oncogenic driver and explain the indolent clinical course of secretory carcinomas. In addition to genetic profiling, identification of mutations has become increasingly relevant in metastatic breast cancer, complementing traditional immunohistochemistry. Mutations of the catalytic subunit α of the phosphatidylinositol 4.5 bisphosphate three kinase gene (PIK3CA), present in up to 40% of luminal tumours, provide eligibility for treatment with the PI3K inhibitor, Alpelisib, while activating HER2 kinase domain mutations are potentially amenable to tyrosine kinase inhibitors such as tucatinib and neratinib. Furthermore, ESR1 (estrogen receptor gene ESR1) mutations contribute to endocrine resistance in a significant proportion of advanced luminal cancers, and NTRK (neurotropic tropomyosin receptor kinase) gene fusions, detected in nearly 50% of secretory carcinomas, predict marked responsiveness to Larotrectinib. These findings underscore the growing clinical importance of predictive molecular pathology in guiding personalised therapy [24].

NTRK fusions, though uncommon, define a therapeutically relevant subset of breast cancers. In a cohort of 23 fusion-positive cases, 11 were secretory (7 with ETV6-NTRK3), 11 non-secretory (7 with NTRK1) and 1 mixed, with secretory tumours occurring in younger, ER-negative patients and non-secretory tumours enriched for TP53 mutations. Querying the TCGA PanCancer Atlas Breast Invasive Carcinoma cohort revealed that none of the 12 male breast cancer samples harboured the ETV6–NTRK3 fusion, while only two female BRCA cases exhibited this rearrangement. These results establish that NTRK fusions extend beyond secretory breast cancer and highlight the critical role of comprehensive genomic profiling in identifying patients eligible for TRK inhibitor therapy [24].

At present, surgery is considered the mainstay of treatment for SBC and can include wide local excision, simple mastectomy, and modified radical mastectomy. We present a rare pediatric male case of metastatic SBC with ETV6-NTRK3 fusion and post-treatment elevated HER2 expression, exploring the implications of precision oncology and the challenges associated with rare malignancies. Detailed medical history has been withheld to preserve patient confidentiality; additional information is available upon request.

### Molecular Methods

RNA and DNA were isolated from the retrospective biopsy specimen from 2023, using TRIzol reagent (#15,596,018; Invitrogen, USA) following manufacturer’s protocol. Briefly, for RNA isolation, tissue was homogenised in TRIzol, 200μl of chloroform (#496,189; Sigma) was added to separate the aqueous phase, and total RNA was precipitated with 100% isopropanol (#DB4DF64078; Merck). The supernatant was discarded, and the pellet was washed with 75% ice-cold ethanol (#MB228; HiMedia). The air-dried pellet was then dissolved in nuclease-free water for further processing. For DNA isolation, genomic DNA was recovered from the interphase and organic phase of the same extraction, precipitated using ethanol, and washed before resuspension. RNA was quantified using Qubit RNA Assay BR (#Q10210; Invitrogen) for sequencing.

Messenger RNA (mRNA) isolated from the biopsy specimen was sequenced on Illumina HiSeq X platform using 150 bp paired-end chemistry. Raw reads were quality-checked and filtered using fastp [25] and subsequently aligned to the human reference genome (GRCh38) using the STAR aligner [26]. Gene expression was quantified as transcripts per million (TPM). Gene fusion detection was performed using STAR-Fusion pipeline on aligned data. Candidate fusions were queried in fusion databases, including ChimerDB 4.0 and FusionGDB2, for validation and occurrence in published cancer cohorts. Whole-exome sequencing (WES) was performed on DNA derived from the same sample using the Illumina HiSeq X platform with 150 bp paired-end reads. Reads were aligned to GRCh38 using BWA-MEM, and variant calling was performed using the GATK best practices pipeline. Identified variants were annotated using the COSMIC (Catalogue of Somatic Mutations in Cancer) database.

Statistical Analysis and Visualisation All analyses were performed in R (version 4.2.2), including differential gene expression using DESeq2 R library. Bar plots and volcano plots were generated using the ggplot2 package. No formal statistical testing was performed, given the single-patient nature of this study.

## Results

### Mutational Analyses

Whole-exome sequencing identified 15 exonic somatic mutations with COSMIC annotations, including four recurrently reported in breast cancer: ACAD11, OR6A2, PTPRM, and INSR. Collectively, these genes implicate key oncogenic processes: metabolic rewiring (ACAD11), dysregulated cell signalling and adhesion (PTPRM, INSR), and emerging roles for ectopic GPCRs in tumour biology (OR6A2) while consistent with breast tumourigenesis, the broader set of variants was most frequently observed in lung cancers (27%) across COSMIC **(Figure 1A)**, a finding of particular relevance given the lung’s status as a common metastatic site in breast cancer. This finding aligned with the clinical presentation of our patient, who developed lung metastases. This overlap underscores potential shared molecular vulnerabilities that may facilitate metastatic tropism to the lung and highlights the biological relevance of the mutational landscape in this rare pediatric SBC case.

**Figure 1:**
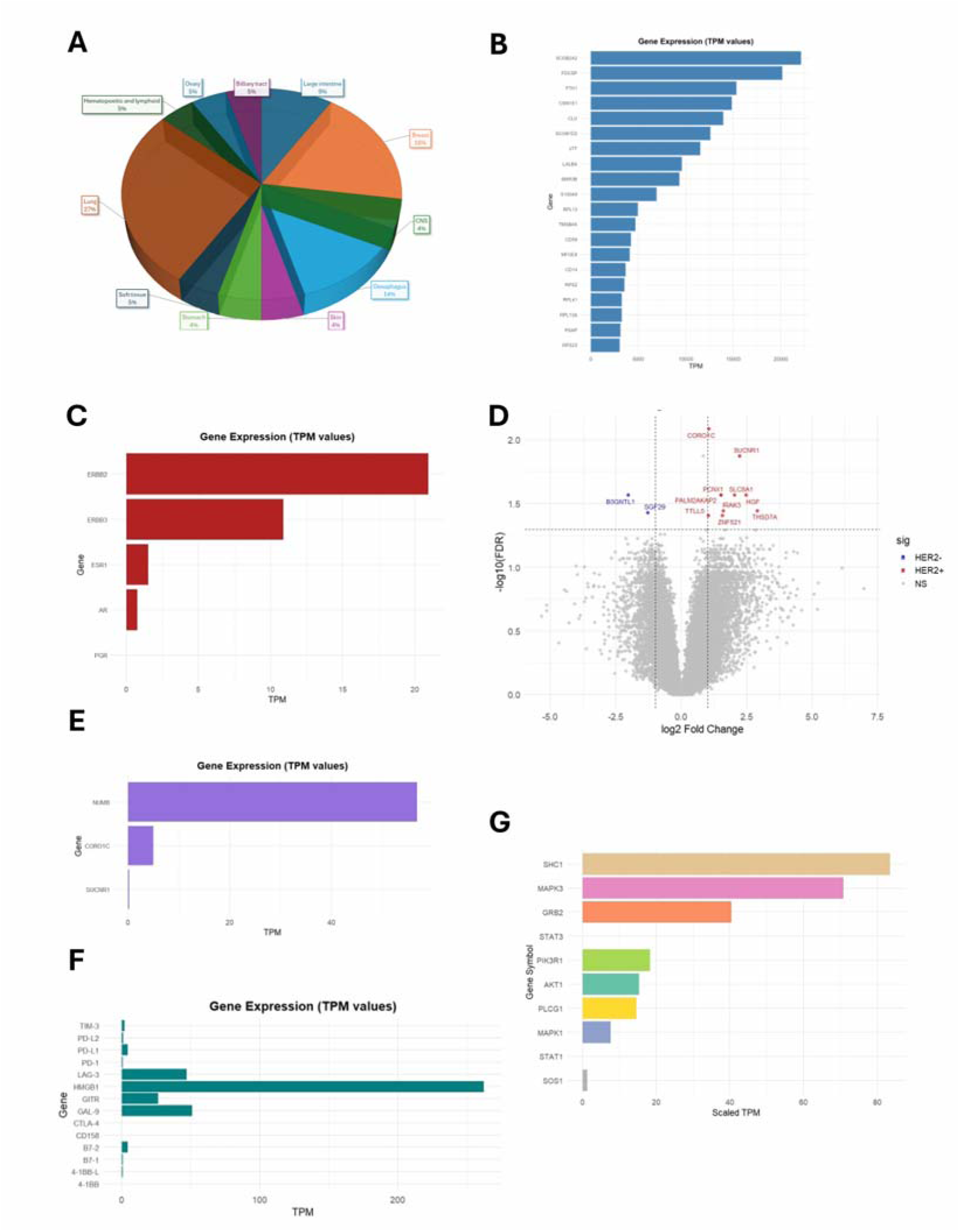
(A) Distribution of 15 exonic mutations identified by WES across cancer types based on COSMIC data, and (B) Normalized TPM values of top 20 highly expressed genes in the patient’s tumour. (C) Expression levels (normalized TPM) of canonical hormone receptors (ESR1, PGR, AR) and HER family receptors (ERBB2/HER2 and ERBB3), (D) Differentially expressed genes in TCGA-BRCA HER2-high and HER2-low MBC tumours (n = 12, stratified by median ERBB2 expression). (E) Expression of HER2-associated genes (CORO1C, NUMB, SUCNR1, among others), (F) Expression of selected immune checkpoint molecules (G) Expression levels of ETV6-NTRK3(e5-e15) variant interactome genes.

### Pathway Analyses

Transcriptomic profiling of the tumour revealed that the most abundantly expressed genes were strongly enriched for secretory and epithelial lineage markers, including SCGB2A2 (mammaglobin-A), SCGB1D2, LTF, LALBA, and CSN1S1. These findings further support the histopathological diagnosis of secretory breast carcinoma, reinforcing the tumour’s mammary epithelial origin despite its unusual clinical behaviour. In addition, highly expressed genes such as FTH1, CLU, and S100A9 are associated with cellular stress responses, iron metabolism, and inflammatory signalling, suggesting an adaptive or pro-tumorigenic microenvironment. The prominence of ribosomal (RPL13, RPS2, RPL41, RPL13A, RPS23) and cytoskeletal (TMSB4X) transcripts reflects the tumour’s high biosynthetic and proliferative activity. The expression landscape highlights the secretory differentiation program and the metabolic–inflammatory adaptations that may contribute to the tumour’s aggressive course. **(Figure 1B)**

### Her2Neu Expression

Furthermore, transcriptome analyses of the tumour revealed absent or negligible expression of canonical hormone receptor genes, including *ESR1* (estrogen receptor alpha), *PGR* (progesterone receptor), and *AR* (androgen receptor), based on scaled TPM values. In contrast, *ERBB2* (HER2) was expressed appreciably, suggesting a HER2-enriched molecular subtype. This expression profile indicates a hormone receptor-negative, HER2-positive tumour, uncommon in secretory breast carcinoma, typically characterised by a triple-negative, indolent phenotype. **(Figure 1C)**.

Differential gene expression (DGE) analysis was performed using RNA-seq data from 12 male patients from the TCGA-BRCA cohort, stratified into groups based on median ERBB2 expression levels to contextualise the tumour’s transcriptomic profile further. **(Figure 1D)**. DGE analysis revealed that the HER2-high tumours exhibited higher levels of genes THSD7A, HGF, SUCNR1, SLC8A1, IRAK3, ZNF521, PCNX1, PALM2-AKAP2, CORO1C, TTLL5 and NUMB compared to tumours in the lower half of HER2 expression. Functional enrichment of these elevated genes converged on negative regulation of protein phosphorylation, suggesting that, alongside HER2-driven hyperactive kinase signalling, a parallel regulatory program may emerge to buffer excessive phosphorylation. HGF and SUCNR1 point toward crosstalk between HER2 and growth factor/GPCR-mediated pathways within this network, whereas NUMB and IRAK3 implicate endocytosis and immune modulation regulatory circuits. On the other hand, the downregulated genes SGF29 and B3GNTL1 in HER2-high tumours, chromatin-associated regulators, suggest suppression of epigenetic modulators in the HER2-high context. *CORO1C* and *NUMB* were also expressed in our patient, providing transcriptomic confirmation of a HER2-enriched subtype. This subtype, characterised by HER2 positivity in the absence of hormone receptor expression, is atypical and relatively uncommon in male breast cancer, where hormone receptor-positive phenotypes are more prevalent. **(Figure 1E)** Though SBC is typically triple-negative, our RNA-based findings suggest HER2-targeted therapy may have therapeutic value even without HER2 protein overexpression.

### Expression of Immune Checkpoint Markers

Next, we evaluated the expression of immune checkpoint markers in the patient’s tumour transcriptome. Among the markers analysed, HMGB1 showed the highest expression with a scaled TPM exceeding 250. Moderate expression levels were observed for LAG3 (∼50 TPM), LGALS9/Galectin-9 (∼50 TPM), GITR, PD-L1 (CD274), and B7-2 (CD86). The presence of these molecules may reflect an immunologically active tumour microenvironment with potential immune evasion mechanisms and may have implications for responsiveness to immune checkpoint blockade therapies. **(Figure 1F)**

### Detection of ETV6–NTRK3 fusion from Transcriptome analysis

The patient was advised to undergo testing for the NTRK3 gene fusion, given the clinical presentation and transcriptomic profile. STAR-Fusion detected multiple putative fusion events; among these, the ETV6–NTRK3 fusion exhibited the highest spanning fragment count (n = 10), strongly supporting its biological relevance, with the breakpoint mapped to exon 5 of ETV6 (chr12:11869969) and exon 15 of NTRK3 (chr15:87940753). This breakpoint combination corresponds to the canonical variant reported in secretory breast carcinoma (SBC) and other tumour types [27]. Further exploration of the fusion interactome revealed strong expression of key adaptor and signalling molecules, notably SHC1 and MAPK3, with moderate expression of PIK3R1, AKT1, PLCG1, and MAPK1. These findings suggest activation of the MAPK/ERK and PI3K/AKT pathways, consistent with the known oncogenic signalling cascade downstream of the ETV6–NTRK3 fusion. **(Figure 1G).** When cross-referenced with ChimerDB 4.0 fusion database, ETV6-NTRK3 fusion with the exact breakpoints was observed in a subset of breast, colon, and skin tumours in the TCGA dataset.

## Discussion

The present work reports a rare case of pediatric male secretory breast carcinoma with triple-negative receptors and pathogenic BRCA2 and MLH1 mutations. Despite prior surgery and systemic therapies, the disease progressed. Transcriptome of the stored post-treatment fresh frozen samples from 2023 identified an actionable ETV6–NTRK3 fusion, and Her2Neu overexpression indicated drug Gemcitabine induced over expression of the marker.

This case emphasises the utility of genomic and transcriptome analysis by next-generation sequencing in uncovering actionable molecular targets in rare and aggressive pediatric SBC. Early genomic and transcriptomic profiling could inform precision therapies such as HER2-directed or TRK inhibitor treatment, enabling tailored management and potentially improving outcomes in ultra-rare, treatment-resistant malignancies.

Her2Neu over expression of a post-treatment sample found to be triple negative at initial presentation indicated therapy-induced upregulation of the marker. Gemcitabine is known to induce HER2 expression through NF-κB signalling [28]. However, methodological variability in IHC of the initial sample can also be a likely explanation. Unlike PD-L1 testing, which benefits from standardised antibody clones and validated scoring systems, HER2 IHC assays employ different antibody clones that target distinct epitopes. Therefore, a false-negative IHC result can arise if the antibody fails to recognise the epitope expressed in the tumour. This underscores the importance of reflex testing, either by fluorescence in situ hybridisation (FISH) or next-generation sequencing (NGS), in cases where HER2 IHC is negative or equivocal typically scored as “2+” by the pathologist.

In this case of pediatric SBC, identifying key exonic mutations provided insights into the unusually aggressive clinical trajectory. While SBC is typically indolent, recurrent mutations observed in both breast and lung cancers, such as those affecting metabolic regulation (ACAD11), cell adhesion and signalling (PTPRM, INSR), and ectopic GPCR biology (OR6A2), raise the possibility that this tumour harboured molecular programs favouring metastatic dissemination. Notably, the lung was the site of metastasis in this patient, consistent with the overlap between the mutational landscape and lung cancer biology. This suggests that the mutational status may have contributed to the tumour’s adaptability, metastatic tropism, and treatment resistance.

Though not a clinically popular method of detection, we reported in this work the ETV6–NTRK3 fusion by whole-exome analysis, done after the patient’s death, for research purposes. ETV6–NTRK3 is a well-established oncogenic driver in secretory breast carcinoma and a targetable alteration for which TRK inhibitors are available. [29] Early identification of the fusion at diagnosis could have enabled a precision-based intervention with an NTRK inhibitor. This case further highlights the systemic barriers faced in low- and middle-income countries, including limited access to advanced diagnostics, interruptions in therapy, and lack of easy availability of targeted drugs such as PARP inhibitors or TRK inhibitors. These challenges significantly delay or prevent the timely implementation of precision oncology in rare and aggressive cancers.

Our findings emphasise a critical point: a single comprehensive NGS assay can often uncover multiple clinically relevant alterations, HER2 expression at the transcript level, actionable exonic mutations, and the structural variants, including gene fusion, that conventional testing may miss. Earlier incorporation of transcriptome analysis by NGS into routine diagnostics could have expedited targeted therapy decisions and altered the clinical trajectory of this patient. We therefore advocate for the systematic integration of transcriptomic testing into the diagnostic and treatment pathway for rare and treatment-resistant cancers. This is the first report to integrate germline predisposition, transcriptomic receptor profiling, differential gene expression, immune checkpoint analysis, and fusion detection in a pediatric male secretory breast carcinoma. These molecular insights expand our understanding of this rare entity and highlight the transformative potential of precision oncology in clinical practice.

## Ethics Statement

This study was conducted in accordance with institutional ethical guidelines. Ethical approval was obtained from the Narayana Health Academic Ethics Committee (Approval No: NHH/AEC-CL-2025-1484). The requirement for informed consent was waived, as the study involved retrospective analysis of anonymized patient data and archival tissue material.

## Data Availability

All data produced in the present study are available upon request to the authors

